# A Preliminary Assessment of Novel Coronavirus (COVID-19) Knowledge and Perceptions in Nigeria

**DOI:** 10.1101/2020.04.11.20061408

**Authors:** P.O. Olapegba, O. Ayandele, S.O. Kolawole, R. Oguntayo, J.C. Gandi, A.L. Dangiwa, I.F.A. Ottu, S.K. Iorfa

## Abstract

This study assessed knowledge and perceptions about COVID-19 among the general public in Nigeria during the initial week of the pandemic lockdown in the country. From March 28 to April 4, 2020, this cross-sectional survey used an anonymous online questionnaire to collect data from respondents within Nigeria. Purposive and snowball sampling techniques were used to recruit 1357 respondents, aged 15-70 years, from 180 cities and towns within Nigeria. Study data were analysed using descriptive statistics. Approximately more than half (57.02%) of the respondents were male with high level of education (48.86% bachelor’s degree or higher). Approximately half of the respondents (46.94%) opined that COVID-19 was “a biological weapon designed by the Chinese government.” About 94% of the respondents identified “contact with airborne droplets via breathing, sneezing, or coughing” as the most common mode of transmission; most respondents associated COVID-19 with coughing (81.13%), shortness of breath (73.47%) and fever (62.79%). “Regular hand washing and social distancing” was selected by most respondents (94.25%) as a way of preventing infection whereas 11.86% reported “consuming gins, garlic, ginger, herbal mixtures and African foods/soups” as preventive measures against COVID-19. Majority of the respondents (91.73%) thought COVID-19 is deadly; and most respondents (84.3%) got 4 or more answers correctly. It was also observed that the traditional media (TV/Radio) are the most common source of health information about COVID-19 (93.5%). Findings revealed that Nigerians have relatively high knowledge, mostly derived from traditional media, about COVID-19. Their perceptions of COVID-19 bear implications across public health initiatives, compliance with precautionary behavior as well as bilateral relations with foreign nations. Evidence-based campaign should be intensified to remove misconceptions and promote precautionary measures.

## 1. Introduction

The novel Coronavirus disease 2019 (COVID-19), first identified in Wuhan China in December 2019, has rapidly spread to almost every region of the world. The disease is caused by a new and severe type of Coronavirus known as severe acute respiratory syndrome coronavirus 2 (SARS-CoV-2). The infection has no immediate treatment and vaccine, and it has according to World Health Organization (WHO, 2020) become a worldwide pandemic causing significant morbidity and mortality. There are 1,603,428 confirmed cases, 356,440 recoveries from the illness and 95,714 deaths worldwide as of April 9, 2020 (Worldometers, 2020). On February 27, 2020, an Italian citizen became the index case for COVID-19 in Nigeria and as at April 9, 2020, there were 288 laboratory-confirmed cases of COVID-19 in Nigeria with 51 discharges and 7 deaths (Nigeria Centre for Disease Control, NCDC, 2020).

To prevent further spread of the virus, civil societies and government agencies embarked on enlightenment campaigns for good hygiene and social distancing. Temperature screening was conducted at airports and those returning from countries with numerous confirmed cases of COVID-19 were implored to self-isolate. The NCDC in association with State governments also began tracing and tracking of possible victims and their contacts. On March 18, 2020, the Lagos State government suspended all gatherings above fifty people for four weeks and ordered all lower and middle level public officers to stay-at-home (Ewodage, 2020). Similarly, the Federal government, on March 30, 2020 introduced various containment strategies such as closing of the national borders and airspace, schools, worship centers and other public places, canceling of mass gathering events and placing the Federal Capital Territory, Lagos and Ogun states on lock down for an initial period of fourteen days (Radio Nigeria, 2020). Covid-19 testing laboratories were set up in Lagos, Abuja and Irrua in Edo State while State governments opened isolation centres and imposed dawn to dust curfews in their territories.

COVID-19, from the family of Coronavirus (others include SARS, H5N1, H1N1 and MERS), is a contagious respiratory illness transmitted through the eyes, nose, and mouth, via droplets from coughs and sneezes, close contact with infected person and contaminated surfaces. It has an incubation period of approximately one to fourteen days. The symptoms include cough, fever and shortness of breath, and it is diagnosed through a laboratory test. The contagion could lead to severe respiratory problems or death, particularly among the elderly and persons with underlying chronic illnesses. Some infected persons however, are carriers for the virus with no symptoms while others may experience only a mild illness and recover easily (Sauer, 2020). As there is currently no cure or vaccine for the COVID-19; medical treatments are limited to supportive measures aimed at relieving symptoms, use of research drugs and therapeutics.

Knowledge of infection pathways and relevant precautions to take is needed to control the pandemic. While the scientific community continues to research possible vaccines or drugs for the viral infection, it is expected that adequate knowledge will motivate individuals to make decisions which may prevent and curb the epidemics. Knowledge such as regular hand washing, using hand sanitizers, wearing face masks, respiratory etiquettes, social distancing and self-isolation when sick are vital to reducing widespread infection (Leppin & Aro, 2009). Studies (e.g. Brug, Aro, Oenema, de Zwart, Richardus & Bishop, 2004; Choi & Yang, 2010; Hussain, Hussain & Hussain 2012) revealed that individuals’ level of knowledge about an infectious disease can make them behave in ways that may prevent infection. Consequently, individuals may need to be informed about the potential risks of infections in order to adopt the right precautionary measures (Brug, Aro & Richardus, 2009).

At early stages of a pandemic, precautionary measures are needed to protect against possible danger and curtail the disease spread. In line with this therefore, the Nigerian government (just like other governments around the world) introduced various containment strategies which have interfered with individuals’ daily lives and have led to severe economic loss and social disruption. People were coerced to stay at home, businesses and offices were closed, exempting healthcare facilities/workers and “essential” commercial establishments. For Nigerians making a living in the informal economy, their livelihood is now threatened by the lockdown since much of their activities and businesses involve face-to-face contact. In Nigeria there is no social safety net, no access to food stamps or unemployment benefits, most people earn their living on a daily basis. Regardless of this however, there has so far been a high degree of compliance with the government directives, Nigerians are engaging in vigilant hand washing, practicing social distancing and self-isolation, and avoiding going to work, school or crowded areas. Even most religious leaders agreed to stop large gatherings, forbid the shaking of hands and directed church members to pray at home and use hand sanitizers (Makinde, Nwogu, Ajaja & Alagbe, 2020; Olatunji, 2020).

On the other hand, some Nigerians due to superstitions and ignorance of the science behind the infection prefer only to pray (even violating the social distancing rule by attending churches or mosques during the lockdown) and use anointing oils, talisman, herbs or rituals (Abati, 2020) to prevent contracting and spreading the virus. Some also use social media platforms (e.g. Whatsapp, Twitter, Facebook and Instagram) to spread fear, project fake news concerning the source of the virus, promote prejudice against China, incite panic buying, proffer fake cures and undermine medical advice, deliberately or ignorantly (Hassan, 2020). They opined that lockdown, self-isolation and social distancing are un-African solutions to the pandemic (Abati, 2020).

Given the importance of knowledge of precautionary activities in curbing the spread of infectious diseases such as the novel COVID-19, it is important to research on people’s health knowledge at this period of the pandemic. Richards (2017) reported that knowledge among ordinary people about how to eliminate risks of contracting Ebola virus led to a rapid drop in mid-2015 in the number of cases of infection. Therefore, in this study, we hope to ascertain the level of the knowledge of COVID-19 among a sample of Nigerians as well as their perceptions of the pandemic. The present study is guided by the following questions:

- What is the knowledge of Nigerians about the source of COVID-19?
- What is the knowledge of Nigerians about the mode of transmission of COVID-19?
- What is the knowledge of Nigerians about the symptoms of COVID-19?
- What is the knowledge of Nigerians on preventive behavior toward COVID-19?
- How fatal do Nigerians perceive COVID-19?
- What are the major sources of information about COVID-19 among Nigerians?

## 2. Methods

### 2.1. Setting and Participants

This cross-sectional survey used an anonymous online questionnaire to collect data from respondents. Potential respondents were purposively sent the link via Social media (Whatsapp and Facebook posts) and asked to participate in an online survey. A snowball sampling technique was employed to recruit more Nigerians living in the country’s six geopolitical zones during the COVID-19 pandemic by encouraging those sent the link to kindly share with their contacts. The online survey ran during the first week of the lockdown in Nigeria (March 28 to April 4, 2020) and involved 1,357 respondents from 180 cities and towns in the country.

### 2.2. Procedure

Due to the Nigerian Government social distance rules and curfew/lockdown enforcement, physical interaction was not possible, so online promotion of the survey was done and existing study participants were urged to send the web link of the survey to potential respondents. They completed the questionnaires hosted on Google online survey platform. Ethical approval was obtained from the Faculty of Social Sciences Ethical Board, University of Ibadan. Participation was completely consensual, anonymous and voluntary, and informed consent was obtained from all respondents.

### 2.3. Instruments

Socio-demographic data were elicited from the respondents on variables such as gender, age, marital status, ethnicity, educational qualification, religion, perceived financial situation and present location.

Knowledge about COVID-19 was assessed using five items adapted from the Ebola knowledge scale developed by Rolison and Hanoch (2015). Components of COVID-19 knowledge included the source of COVID-19, modes of transmission, symptoms, methods of preventing and curbing the infection, perception of COVID-19 fatality and sources of information about COVID-19. Respondents’ knowledge about COVID-19 is arrived at by summing correct responses across item 1, source of COVID-19, (correct = [d]), item 2, transmission of COVID-19, (correct = [a], and [b], [c] or [d)), item 3, prevention of COVID-19, (correct = [b] and [d], [f] or [h]), item 4, symptoms of COVID-19, (correct = [a], [b] and [g]), and item 5, awareness of COVID-19 fatality, (correct = [a]), generating a maximum possible score of five. The norm is set at 3 which indicated moderate level of knowledge about COVID-19. Scores above 3 indicated high level of knowledge about COVID-19 while scores less than 3 indicated low level of knowledge about COVID-19. The mean score and standard deviation for the sample population was calculated to indicate the sample’s level of knowledge. Similarly, scores above the norm indicated high knowledge and score below the norm indicated low knowledge of COVID-19 for the sample.

### 2.4. Statistical Analysis

Descriptive statistics was used for respondents’ socio-demographic characteristics and knowledge about COVID-19. Percentages of response were calculated according to the number of respondents per response. Descriptive statistical analysis was performed using IBM SPSS Statistic version 20.

## 3. Results

### 3.1. Survey Respondents

We received responses from 1403 respondents, as at 4th April, 2020, which was the data cutoff collection date for this study. We included 1357 respondents from 180 cities and towns in Nigeria who had completed the online questionnaires (completion rate: 95.72%). The respondents were aged between 15-70 years (Mean 26.85, SD = 9.17). There were 570 female respondents (42.0%) and almost all respondents (97.5%) had completed at least high school education. Majority (68.46%) were Yoruba in South west Nigeria, the region with the highest reported cases of COVID-19 infection (NCDC, 2020). Table 1 provides the sample demographics.

**Table 1.** Sample characteristics (n = 1,357) Age (Mean 26.85, SD = 9.17)

| Variables | Categories | Frequency | Percent |
| --- | --- | --- | --- |
| Gender | Female | 570 | 42.00 |
|  | Male | 787 | 58.00 |
| Educational qualification | High School | 349 | 25.72 |
|  | Diploma | 313 | 23.07 |
|  | Degree | 421 | 31.02 |
|  | Higher degree | 242 | 17.83 |
|  | Others | 32 | 2.36 |
| Perceived Financial Status | Don't meet basic needs | 246 | 18.13 |
|  | Just meet basic needs | 479 | 35.30 |
|  | Meet needs with a little left | 387 | 28.52 |
|  | Live comfortably | 245 | 18.05 |
| Relationship Status | Single/Not dating | 560 | 41.27 |
|  | Single/but dating | 524 | 38.61 |
|  | Married | 269 | 19.82 |
|  | Separated/divorced/widowed | 4 | 0.29 |
| Ethnic Grouping | Hausa-Fulani | 138 | 10.17 |
|  | Igbo | 115 | 8.47 |
|  | Yoruba | 929 | 68.46 |
|  | Other ethnicities | 175 | 12.90 |
| Religion | Christianity | 842 | 62.05 |
|  | Islam | 506 | 37.29 |
|  | Others | 9 | 0.66 |

### 3.2. Knowledge of COVID-19

What is the knowledge of Nigerians about the source of COVID-19?

Approximately half of the respondents (46.94%) opined that COVID-19 is “a biological weapon designed by the government of China” while 41.93% identified it as “a severe illness transmitted to people from wild animals” (fig. 1).

**Fig. 1:**
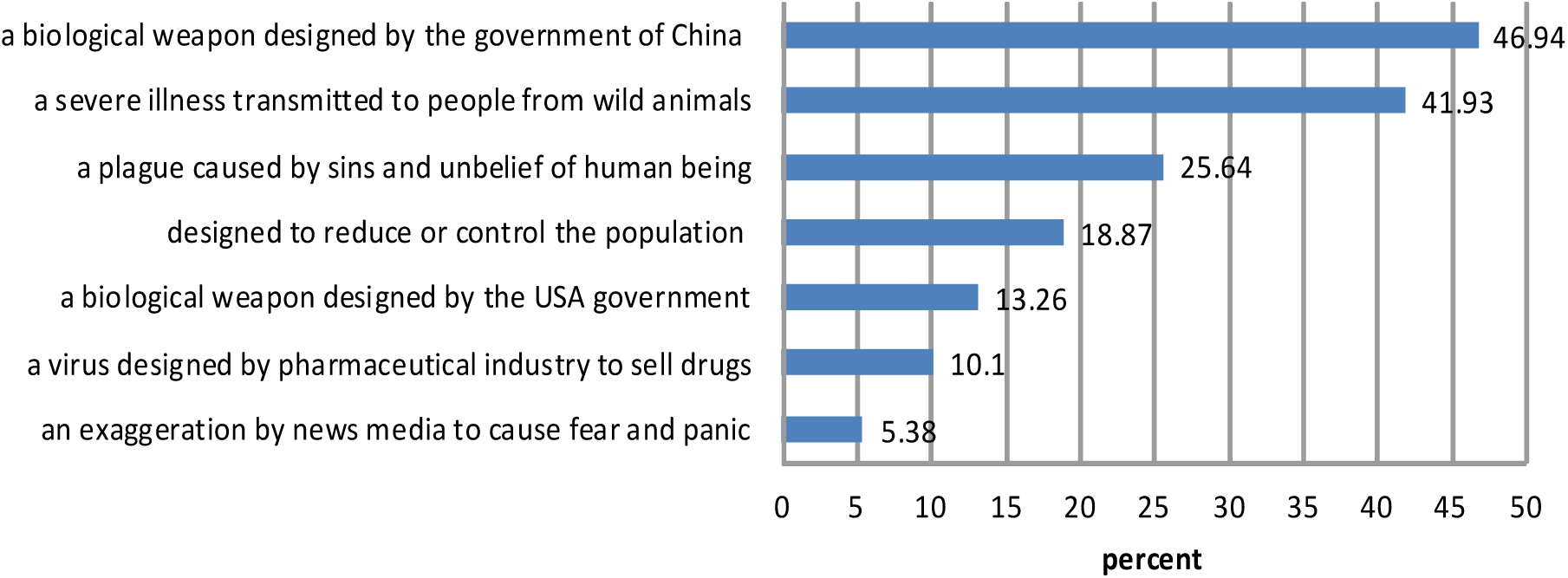
Sources of COVID-19

What is the knowledge of Nigerians about the mode of transmission of COVID-19?

Regarding knowledge about the most common perceived mode of transmission, almost all (94.10%) selected “contact with droplets from an infected person/organism via breathing, sneezing, or coughing” while a little above average (54.97%) picked “touching contaminated objects or surfaces” as a mean of transmitting and contacting the virus (fig. 2).

**Fig. 2:**
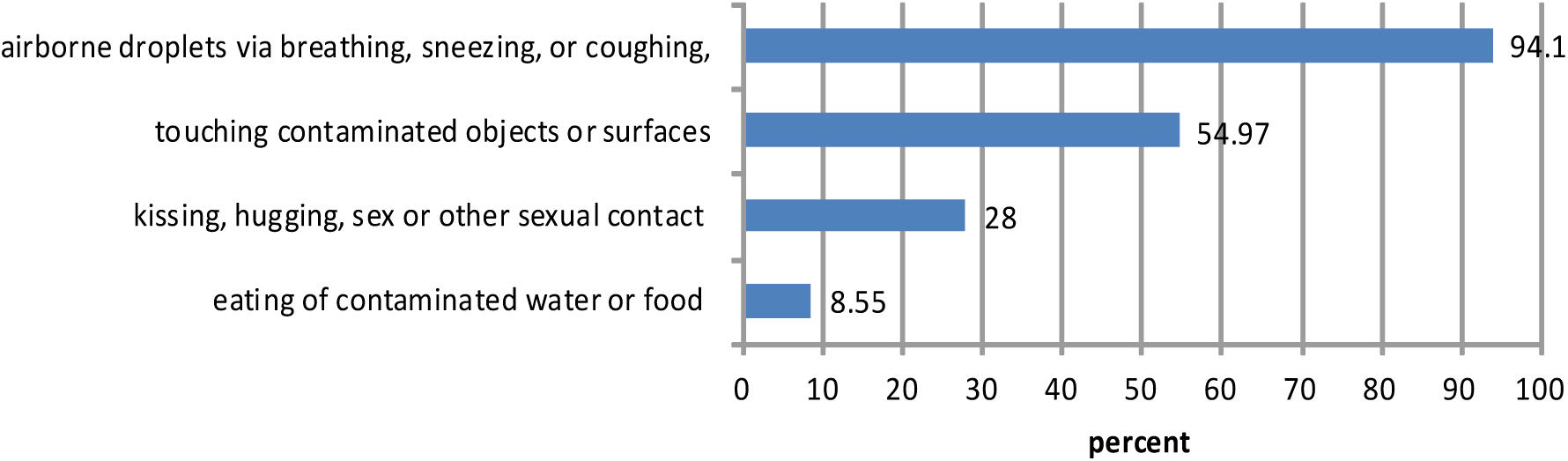
Transmission of COVID-19

What is the knowledge of Nigerians about the symptoms of COVID-19?

Most respondents accurately associated COVID-19 with coughing (81.13%), shortness of breath (73.47%), sneezing (69.71%), fever (62.79%) and sore throat (56.82%) while 1.62% claimed to have no idea about the symptoms of the disease (fig. 3).

**Fig. 3:**
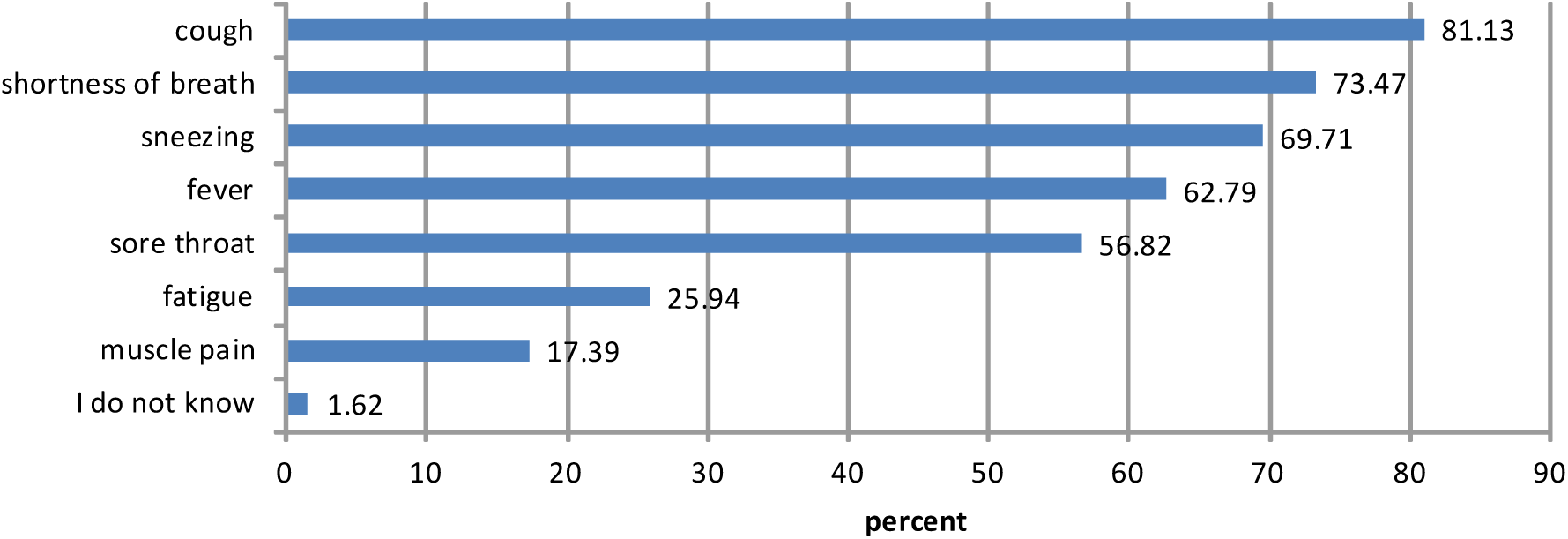
Symptoms of COVID-19

What is the knowledge of Nigerians on preventive behavior toward COVID-19?

“Regular hand washing and social distancing” were selected by most respondents (94.25%) as a way of preventing COVID-19 infection while about half (48.86%) supported “disinfecting contaminated surfaces” and 40.01% supported “closing schools and cancelling mass gathering events” whereas more than a tenth (11.86%) held “consuming gins, garlic, ginger, herbal mixtures and African foods/soups” as preventive measures against COVID-19 (fig. 4).

**Fig. 4:**
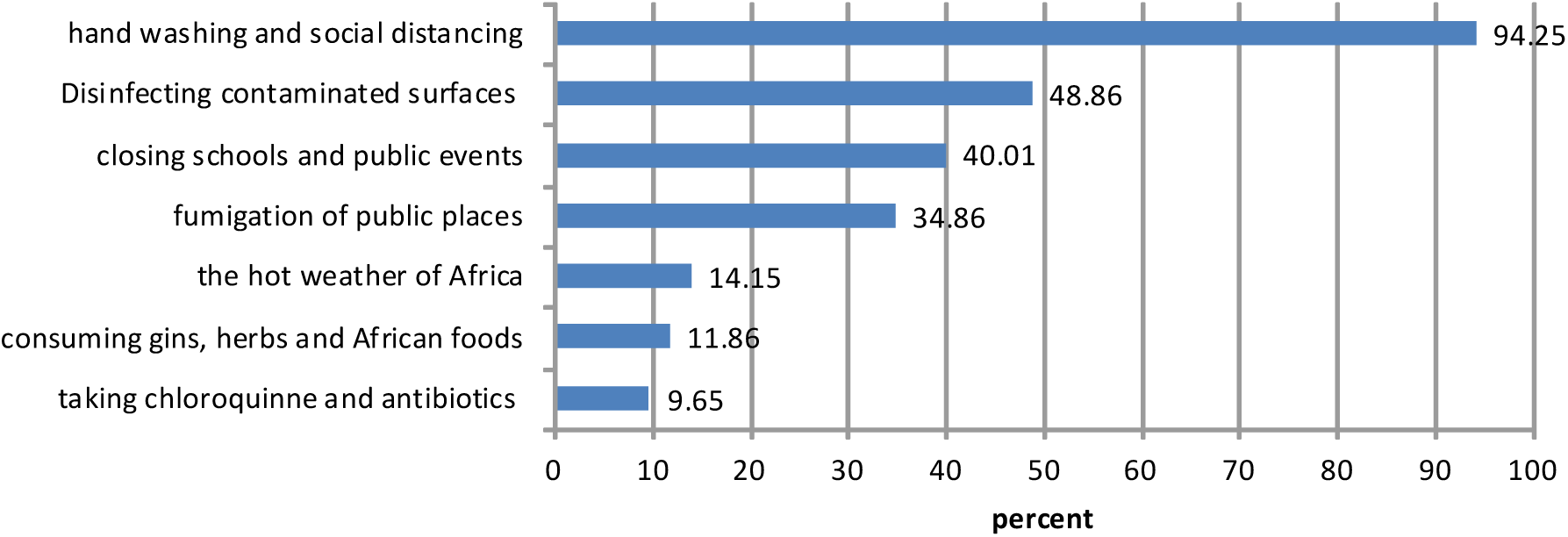
Prevention of COVID-19

How fatal do Nigerians perceive COVID-19?

Fig. 5 revealed that majority of the respondents (91.73%) acknowledged that “it is possible to die from COVID-19.”

**Fig. 5:**
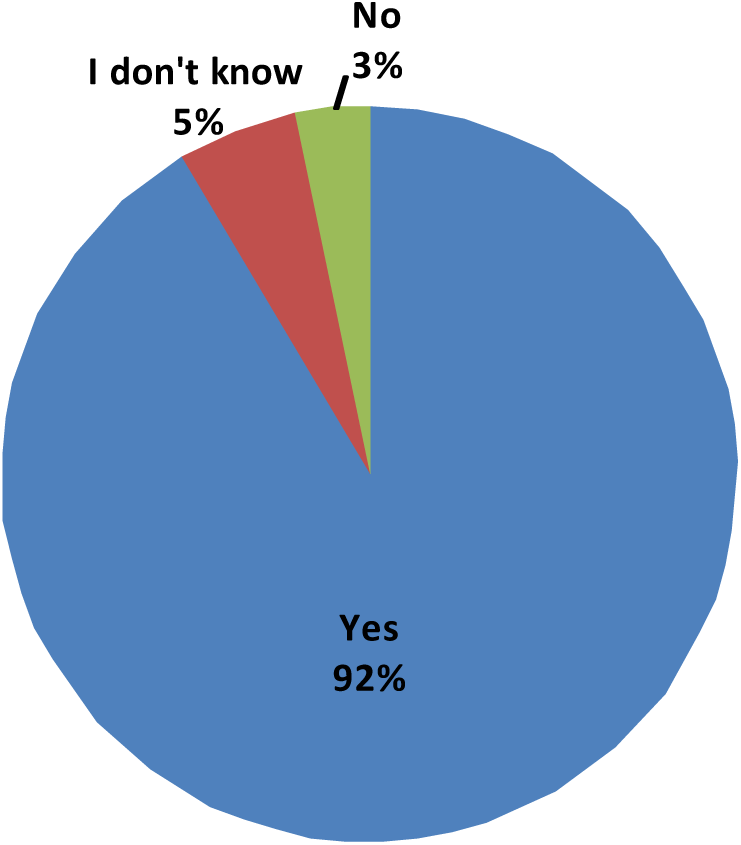
Possible death from COVID-19 infection

What are the major sources of information about COVID-19 among Nigerians?

The most common sources of information about COVID-19 were the mass media (radio, television and newspapers) (81.5%) followed by social media (e.g. Whatsapp, Facebook, Twitter, Instagram, etc.) and the Internet (61.53%) (fig. 6).

**Fig. 6:**
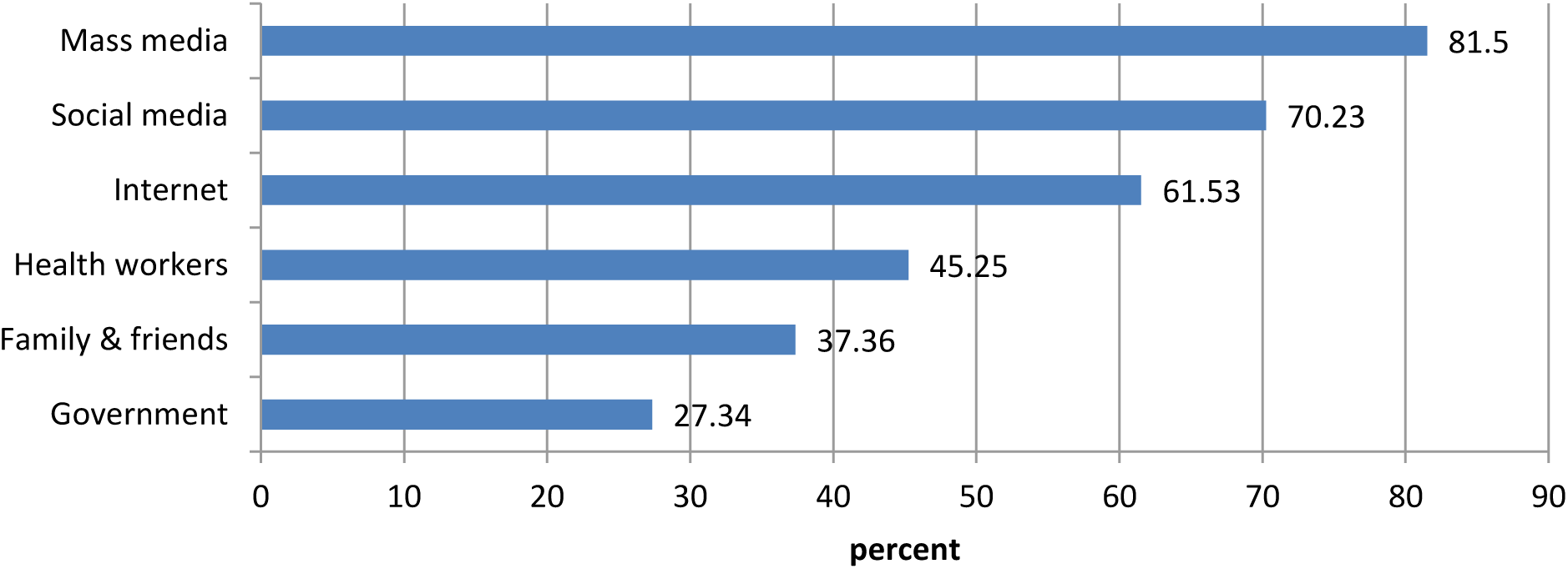
Sources of Information about COVID-19

Table 2 below showed that most respondents (83.86%) had high knowledge about COVID-19 (got more than 3 answers correctly) and 13.34% reported average knowledge (3 correct answers) while 2.79% had low knowledge (less than 3 correct answers) about COVID-19. Respondents in this sample had a mean total knowledge score of 4.17 (SD = 0.77) which is above the norm score of 3.

**Table 2:** Total knowledge score.

| Correct answers | Frequency | Percent | Mean (SD) |
| --- | --- | --- | --- |
| None | 1 | 0.07 | 4.15 (0.77) |
| One | 4 | 0.29 |  |
| Two | 33 | 2.43 |  |
| Three | 181 | 13.34 |  |
| Four | 667 | 49.15 |  |
| Five | 471 | 34.71 |  |

## 4. Discussion

Since its discovery in 2019, the COVID-19 seems to have become one of the largest pandemics in the world involving more than 200 countries (Worldometers, 2020). This study set out to assess the knowledge of the general public in Nigeria about COVID-19 during the initial week of the pandemic lockdown in the country. For individuals to survive in the era of pandemic, adequate knowledge of the disease that caused the pandemic is required. Such knowledge can help to contain the pandemic by adopting right precautionary measures, which will invariably boost both the physical and mental health of the individuals.

Findings from this study indicated that a large proportion of the study participants are aware and knowledgeable about the COVID-19 and its presence in Nigeria. Results obtained from the research questions regarding knowledge of COVID-19 in terms of respondents’ knowledge of the source of COVID-19, transmission of COVID-19, symptoms of COVID-19, preventive behavior toward COVID-19, fatality rate of the COVID-19 and what the major sources of information about COVID-19 among Nigerians are, were significantly high.

Specifically, this study found that a large percentage of Nigerians hold the view that the COVID-19 is a biological weapon designed by the government of China. This is evident of the diverse sources of information concerning the COVID-19 that is available to Nigerians (Hassan, 2020). What this means is that there may currently be no consensus among Nigerians as to what the real source of the virus is. We caution that this perception has implications for bilateral relations between the Nigerian and the Chinese governments and may stand as a hindrance to Nigerians accepting whatever form of aid may come from the Chinese government. It is important the government of Nigeria and other stake holders embark on campaigns to raise awareness of the true sources of the COVID-19 in order to curb a brewing stereotype and prejudice towards the Chinese.

Concerning the source of the COVID-19 also, our findings highlight implications for religious leaders. A reasonable percentage of Nigerians also opined that the COVID-19 is a plague caused by sins and unbelief of human beings. While this may be consistent with many religious beliefs, we believe that it may foster carefree attitudes in Nigerians, making them relax and resort to only prayers and spiritual healings without adhering to the prescribed hygiene practices (Abati, 2020). We therefore urge clerics at all levels to also educate members of their faiths about the COVID-19.

As expected, because Nigerians had relatively high knowledge of the COVID-19, even though laden with several misconceptions, their knowledge of precautionary behavior was also high. For instance, a good percentage agreed that a range of WHO approved and global practices such as hand washing and social distancing, disinfecting contaminated surfaces, closing schools and public events and fumigation of public places were key to preventing the spread of the virus. Only a little percentage agreed relying on the hot weather in Africa, consumption of gins, herbs and African foods as well as chloroquine and antibiotics as precautions to the spread of the pandemic.

These findings are in consonance with previous studies such as Brug et al. (2004), Choi and Yang (2010) and Hussain et al. (2012). These authors reported that one’s level of knowledge about an infectious disease can make one to behave in ways that can prevent infection. Also, the study supported Richards (2017) who opined that knowledge among ordinary people about how to eliminate risks of contracting Ebola virus led to a rapid drop in mid-2015 in the number of cases of infection. Consequently, individuals need to be informed about the potential risk of infection (COVID-19) in order to adopt the right precautionary measures. One suggestion for this result is the knowledge factor. That is, Nigerians have knowledge of COVID-19 and therefore are better able to adopt precautionary measures.

Majority of the respondents (more than 90%) agreed that COVID-19 has high fatality and this is confirmed by the reported 79,384 deaths worldwide as of April 7, 2020 (Worldometers, 2020).

The findings also identified the mass media as the major sources of information about COVID-19 which is similar to a study conducted during the SARS epidemic in Hong Kong (Lau, Yang, Tsui, & Kim, 2003). Likewise Vartti, Oenema, Schreck, Uutela, de Zwart, Brug, and Aro (2009) and Voeten, de Zwart, Veldhuijzen, Yuen, Jiang, Elam and Brug (2009) confirmed that the traditional media provide vital information during outbreaks. However, it contradicted Rolison and Hanoch (2015) which revealed that the internet is the premier source of knowledge during an outbreak. The media should be intensively used by governmental and non-governmental agencies to provide regular enlightenment on proper social distancing, correct personal hygiene and usage of personal protective equipment to ensure compliance with the WHO approved strategies for curbing the pandemic.

### Limitations

The time-sensitivity of the novel Coronavirus disease 2019 pandemic led to the adoption of the snowball sampling strategy which might limit generalizability of the finding to the general population. There was an oversampling of respondents from the Yoruba ethnic group, leading to selection bias. Similarly the findings may not be generalizable to the less educated people. Notwithstanding the above limitations, this study provides a baseline of information on knowledge and perceptions about the ravaging COVID-19 from respondents across 180 municipalities in Nigeria. It is worthy of note that this study is an exploratory one and is part of a larger study aimed at understanding and outlining how knowledge and awareness of COVID-19 among Nigerians is shaping their response to the pandemic and precautionary behavior. There is need for further research to build the evidence base for the study of COVID-19 knowledge and precautionary measures.

## 5. Conclusions

Due to the concern of everyone about COVID-19, the present study, in possibly, is the first survey to assess the knowledge and perceptions about COVID-19 among Nigerians. This study was significant because it studied knowledge about COVID-19 in Nigeria across different and many cities in the country. The knowledge areas include source, transmission, symptoms, sources of information and preventive behavior toward COVID-19. The findings tentatively affirm that Nigerians are highly knowledgeable about COVID-19and their premier sources of information about the pandemic are the traditional media. It is therefore recommended that all stake holders should intensify their effort in sensitizing the general public to understand and comply with all precautionary measures to curb COVID-19.

## Data Availability

Data are available

## Author Contributions

Study conception and design: POO, OA, SKI

Acquisition of data: POO, OA, SOK, RO, ALD, JCG, SKI, IFAO

Analysis and interpretation of data: OA, RO

Drafting of manuscript: OA, SOK, SKI

Critical revision: POO, IFAO, JCG

All authors read and approved the final manuscript for publication

## Questionnaire

Section A: Socio-demographics

1. Age____ years
2. Gender: Male [ ] Female [ ]
3. Relationship status: Single/Not dating [ ] Single/but seriously dating [ ] Married [ ] Divorced/separated/widowed [ ]
4. Ethnicity:________________________
5. Present location:________________________
6. Religion: Christianity [ ] Islam [ ] Traditional [ ] Others [ ]
7. Considering your own income and the income from any other people who help you, how would you describe your overall personal financial situation? don’t meet basic needs [ ] just meet basic needs [ ] meet needs with a little left [ ] live comfortably [ ] Section B: Knowledge of Coronavirus
8. To the best of your knowledge, the novel Coronavirus is: (You may choose more than one).
  a. a biological weapon designed by the government of China
  b. a virus designed by pharmaceutical industry to sell their drugs
  c. an exaggeration by news media to cause fear and panic
  d. a severe illness transmitted to people from wild animals
  e. a plague caused by sins and unbelief of human being
  f. designed to reduce or control the population
  g. a biological weapon designed by the USA government
9. The Coronavirus is typically spread (i.e., passed from person-to-person) by which means? (You may choose more than one).
  a. contact with airborne droplets via breathing, sneezing, or coughing,
  b. kissing, hugging, sex or other sexual contact
  c. eating of contaminated water or food
  d. touching contaminated objects or surfaces
  e. through 5G phone network or masts
  f. using the test-kits or vaccine
  g. living a sinful life
10. Coronavirus can be prevented by
  a. the hot weather of Africa
  b. regular hand washing and social distancing
  c. taking chloroquinne capsules and antibiotics
  d. fumigation and spraying bus stops and other public places
  e. consuming gins, garlic, ginger, herbal mixtures and traditional food and soup
  f. closing schools and cancelling mass gathering events
  g. disinfecting contaminated surfaces
  h. anointing oil and prayers
11. The most important symptoms of COVID19/Coronavirus are: (You may choose more than one).
  a. cough
  b. fever
  c. fatigue
  d. sneezing
  e. sore throat
  f. muscle pain
  g. shortness of breath
  h. I do not know any symptoms of COVID19/Coronavirus
12. Do you think it is possible to die from the Coronavirus? (a) yes, (b) no, and (c) I do not know
13. Which of the following sources have you received new information about the Coronavirus? (You may choose more than one).
  a. The mass media (television, newspapers, radio etc.)
  b. the internet (Google, Wikipedia, etc.)
  c. Health workers (doctors, nurses, pharmacist, NCDC, etc.)
  d. Government officials (governors, ministers, commissioners, etc.)
  e. Friends and family members
  f. Social media (Whatsapp, Facebook, Instagram, Twitter etc.)
  g. other: …

